# Sensory Impairments and Social Isolation Among Hispanic Older Adults: Towards Culturally Sensitive Measurement of Social Isolation

**DOI:** 10.1101/2021.11.16.21266422

**Authors:** Corinna Trujillo Tanner, Jeremy B. Yorgason, Stephanie Richardson, Alisha H. Redelfs, Melanie M. Y. Serrao Hill, Avalon White, Kyriakos S. Markides, Brian Stagg, Joshua R. Ehrlich

## Abstract

**Objectives:** Sensory disabilities, including vision disability and hearing disability, increase risk for social isolation, which is associated with multiple negative health outcomes. Existing literature suggests that the cultural value of familism may provide a buffer against social isolation. We examined the longitudinal trajectory of social isolation among Hispanic older adults with self-reported vision disability (SRVD) and self-reported hearing disability (SRHD) and tested a modified measure of social isolation incorporating familism.

**Methods:** We compared 8-year trajectories of social isolation among Hispanics (*n* = 445) and non-Hispanic Whites (*n* = 4,861) from the National Health and Aging Trends Study. We used structural equation modeling to explore the longitudinal relationships between sensory disability and social isolation while comparing two measures of social isolation.

**Results:** Social isolation increased longitudinally for both groups, with SRVD significantly associated with higher initial levels. Social isolation started and remained higher across time among Hispanics. Using an adjusted measure of social isolation (added familial support), neither initial levels nor trajectories of social isolation differed between Hispanic and non-Hispanic White participants.

**Discussion:** Initially, Hispanics appeared more socially isolated, reporting less social support from outside the home. Yet, we found that they were more likely to report family social connections. Traditional measures of social isolation focusing on social support outside of the home (neglecting support by family) may lack content validity among Hispanic groups. Culturally sensitive measures of social isolation will be increasingly consequential for future research and health policy to meet the needs of a diverse older population.

## Introduction

Cultural diversity in the United States (U.S.) is enhanced by its largest ethnic group, Hispanics, who comprise 18% of the population. The term *Hispanic* describes a heterogeneous group of people who trace their heritage to Latin America or Spain. Individuals who identify as Hispanic may be of any race. In 2019, the Hispanic population in the U.S. was 60.6 million, a 20% increase from 2010 (U.S. Census Report, 2020). Contributing to a trend of increasing ethnic diversity in the U.S., this number is projected to increase to 118 million by the year 2060 when Hispanics will comprise approximately 28% of the U.S. population. Consistent with the aging of the U.S. population, the median age of Hispanics in the country is also increasing (Noe-Bustamante et al., 2020).

Despite great diversity among Hispanics, this group may share common cultural values, which some have suggested contribute to a sociocultural health advantage, referred to as the *Hispanic Paradox*. Although they may, on average, experience higher poverty rates and less access to education, health care, and other resources, Hispanics have a higher life expectancy and better health outcomes in several domains compared with non-Hispanic Whites or Blacks (Markides & Eschbach, 2005). This is especially true among immigrants, although the health advantage seems to equalize in older age (Angel, 2009; Markides & Rote, 2019).

Family-centered values, or *familism*, foster strong social cohesion and sturdy social networks (Gallo et al., 2009; Ruiz et al., 2016). Familism is described as “a cultural frame of reference about the centrality of the family that is enacted in attitudes and behaviors” (Hernandez, 2016, p. 464) u and explains how Hispanics are likely to refer to the family for support, comfort, and services (Behnke, 2008). Related constructs noted in Hispanic culture include filial piety and, *respeto*, which involve honoring family, including caring for aging parents. These communalistic values prioritize social relationships with extended family networks over individual achievement, and create social support via a tight social network that buffers stress (Corona et al., 2017) and promotes resilience (Ruiz et al., 2016). Benefits of strong family social connections may partly explain the longevity and resilience described by the Hispanic Paradox (Markides et al., 2013). Familism seems to be a feature Hispanic ethnicity, regardless of country of origin (Campos et al., 2014)

An important health related outcome associated with sensory impairments is social isolation. Older adults who experience vision impairment, or hearing loss are at significantly increased risk of becoming socially isolated (Shah et al., 2020; Shukla et al., 2020). Social isolation has been linked to a host of negative outcomes, including higher likelihood of anxiety and depression (Domènech-Abella et al., 2019; Santini et al., 2020), worse cognitive functioning (Evans et al., 2019; Read et al., 2020), decreased physical and mental health (Hawton et al., 2011), and mortality (Holt-Lunstad et al., 2015). Vision loss and hearing loss are prevalent among older adults and are increasing across all demographics as the U.S. population ages (Swenor et al., 2013).

Vision loss affects 9% of older adults in the U.S. (Patel et al., 2020), yet risk may be higher for Hispanics. Vision loss among Hispanics is driven by health disparities which influence incidence and access to treatment of diabetic eye disease and cataracts (Herren & Kohanim, 2016; Varma et al., 2004). The unmet need for refractive correction (regular eye glasses) can cause or compound vision impairment. While up to 64% of Hispanics over age 40 have a need for refractive correction, 20% of those lack access, especially those with lower rates of acculturation, lower education level and those without insurance (Uribe et al., 2011). Though Hispanics are at an increased risk for vision loss, they are less likely to receive screening. Sixty-three percent of the participants in the Los Angeles Latino Eye Study who had vision disabilities had never been diagnosed or sought treatment prior to the study (Varma et al., 2004).

Among those of older age, rates of hearing loss are similar between Hispanic older adults and non-Hispanic White older adults (Cruickshanks et al., 2015) and affects 31% of those age 60 to 69 and 63.1% of those age 70 and older (Goman & Lin, 2016). However, individuals of Hispanic ethnicity can expect to live a greater proportion of their lives hearing impaired than non-Hispanics (West & Scott, 2021). Previous research has identified underuse of hearing aids among Hispanic older adults compared with non-Hispanic whites, largely due to lack of health insurance access (Arnold et al., 2019). This suggests a higher impact of hearing loss among Hispanics.

Together, these patterns suggest a strong impact of sensory disabilities for Hispanic older adults, who have less access to resources that mitigate the challenges created by sensory loss, including early diagnosis and treatment and access to adaptive or corrective equipment. Yet, little is known about the possible buffering effect of family-centered values on the myriad adverse health outcomes associated with sensory disability.

Although some research has examined how social isolation differs among older adults based on ethnic origins (Locher et al., 2005), little is known about the direct influence of culture. There is evidence that Hispanic older adults prefer to receive social support primarily from family systems rather than from the community at large (Min & Barrio, 2009). This may be due in part to the cultural value of familism. While established measures of social isolation (Lubben, 1988; Zimet et al., 1988) include measures of family support, researchers who use large, population-based data sets, such as the national health and aging trends study (NHATS), must create measures from existing data and are somewhat limited. Though care may be taken in selecting items to include, they may not be appropriate for individuals of diverse backgrounds. For example, using the National Health and Aging Trends Study (NHATS), the study by Cudjoe et al. (2020) uses a composite measure of social isolation which may underestimate the scope of social support experienced by Hispanic older adults. The typology of this scale relies on social supports that are external to the family, including volunteering in the community and participating in clubs or classes. Other studies utilizing this typology like those by Suntai & White (2021) and Falvey et al. (2021) may be overestimating social isolation among Hispanics as they are not measuring support form family.

Marín and Marín (1991) cautioned that measures may represent the world view of those doing the research and may therefore be culturally biased. Items included may be selected based on the researcher’s perceptions, norms, and values and may lack the ability to reflect the cultural assumptions underlying the respondents’ views.

As the U.S. is becoming more ethnically diverse, understanding the impact of sensory disabilities and social isolation among Hispanic people will be increasingly consequential to future research and policy as we work to understand the intersection of “ethnicity, aging and health” (Howard, 2019, p. 3). Thus, the purpose of this research was to explore the longitudinal trajectory of social isolation among Hispanic older adults with self-reported sensory disabilities, including Self-Reported Vision Disability (SRVD) and Self-Reported Hearing Disability (SRHD). A comparison with non-Hispanic Whites provided a reference point for understanding features of social isolation that may be unique to the Hispanic population. In this process, we explored elements of a more culturally sensitive measure of social isolation in this group, including items addressing familism.

The following questions guided the research:

1. Do trajectories of social isolation differ between Hispanic and non-Hispanic White respondents?
2. Do trajectories of social isolation differ for Hispanic and non-Hispanic White respondents when a more culturally sensitive measure of social isolation is used?
3. Do sensory disabilities relate to trajectories of social isolation differently for Hispanic and non-Hispanic White respondents?

## Methods

### Sample and Procedures

This study used data from the NHATS, a nationally representative longitudinal study of Medicare beneficiaries aged 65 and older. Annual data collection began in 2011, and the current analysis used data from the 2011 cohort followed through 2018 (Rounds 1 through 8). At Round 1, there were initially a total of 8,245 participants in the sample. However, 2,939 participants were dropped for either (1) not living in a community setting or (2) not belonging to the ethnic or racial groups of Hispanic or White non-Hispanic, or missing on either of those characteristics. The resulting analytic sample included 5,306 participants, among whom 4,861 were non-Hispanic White and 445 were Hispanic. The sample did experience attrition over the 8 years of the study. Attrition was consistently around 15% between each wave of the study, with the sample dropping to 32% of the original sample at Round 8.

As seen in Table 1, descriptive statistics indicated an even distribution of participants across age, sex, education, and income. Across the total sample, participants ranged in age from 65 to ≥95, with approximately 20% of the sample falling in each age category (65–69, 70–74, 75–79, 80–84, 85+). Approximately 56% of participants were female, and nearly half (48%) had at least some education post-high school. The average income in the sample was as would be anticipated, yet had considerable variability (Mean = $56,000/year, *SD* $199,383).

**Table 1.**
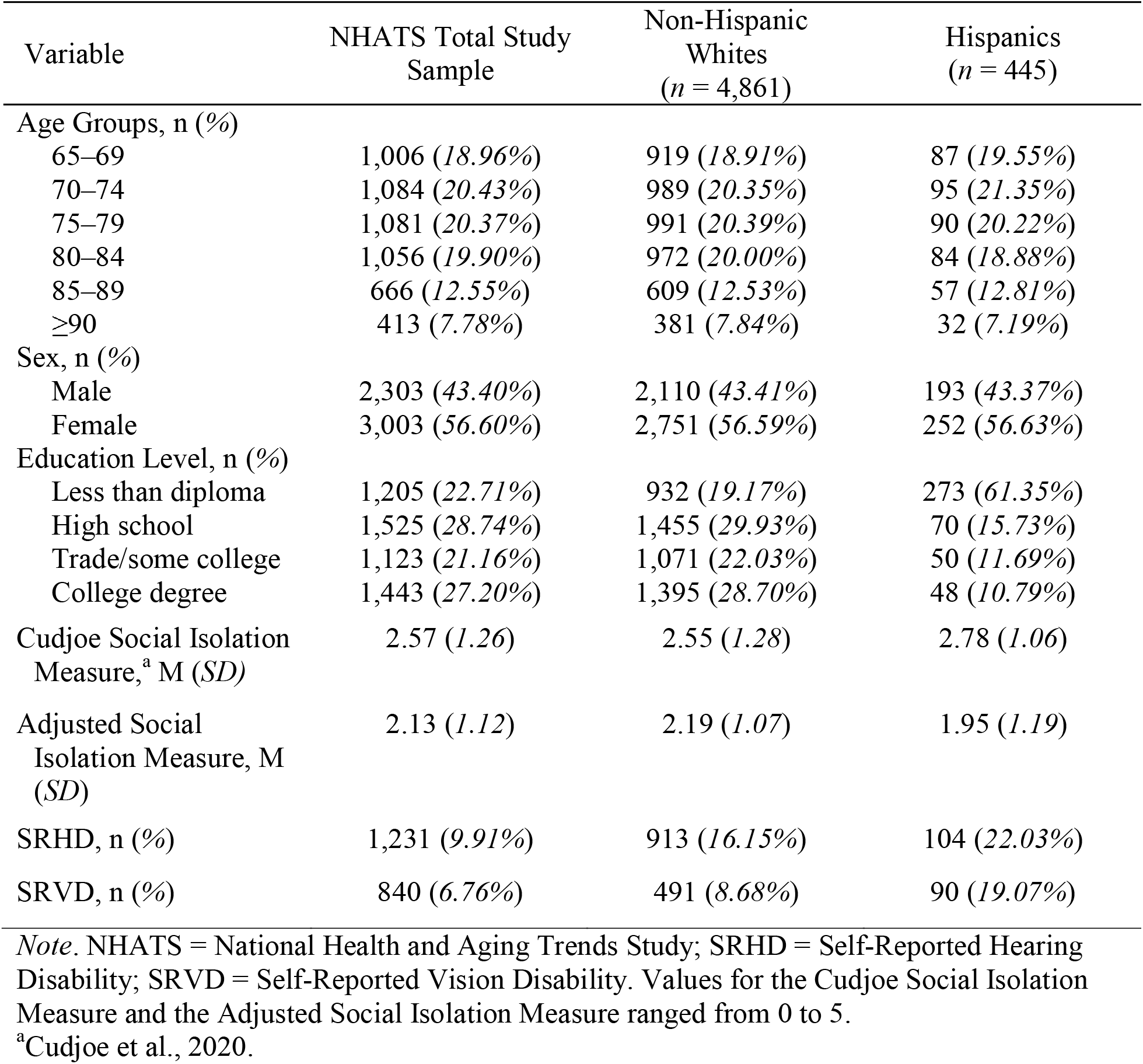
Round 1 of NHATS: Main Study Variables and Demographics

| Variable | NHATS Total Study Sample | Non-Hispanic Whites<br>(*n* = 4,861) | Hispanics<br>(*n* = 445) |
| --- | --- | --- | --- |
| Age Groups, *n* (%) |  |  |  |
| 65–69 | 1,006 (18.96%) | 919 (18.91%) | 87 (19.55%) |
| 70–74 | 1,084 (20.43%) | 989 (20.35%) | 95 (21.35%) |
| 75–79 | 1,081 (20.37%) | 991 (20.39%) | 90 (20.22%) |
| 80–84 | 1,056 (19.90%) | 972 (20.00%) | 84 (18.88%) |
| 85–89 | 666 (12.55%) | 609 (12.53%) | 57 (12.81%) |
| ≥90 | 413 (7.78%) | 381 (7.84%) | 32 (7.19%) |
| Sex, *n* (%) |  |  |  |
| Male | 2,303 (43.40%) | 2,110 (43.41%) | 193 (43.37%) |
| Female | 3,003 (56.60%) | 2,751 (56.59%) | 252 (56.63%) |
| Education Level, *n* (%) |  |  |  |
| Less than diploma | 1,205 (22.71%) | 932 (19.17%) | 273 (61.35%) |
| High school | 1,525 (28.74%) | 1,455 (29.93%) | 70 (15.73%) |
| Trade/some college | 1,123 (21.16%) | 1,071 (22.03%) | 50 (11.69%) |
| College degree | 1,443 (27.20%) | 1,395 (28.70%) | 48 (10.79%) |
| Cudjoe Social Isolation Measure,<sup>a</sup> *M* (*SD*) | 2.57 (1.26) | 2.55 (1.28) | 2.78 (1.06) |
| Adjusted Social Isolation Measure, *M* (*SD*) | 2.13 (1.12) | 2.19 (1.07) | 1.95 (1.19) |
| SRHD, *n* (%) | 1,231 (9.91%) | 913 (16.15%) | 104 (22.03%) |
| SRVD, *n* (%) | 840 (6.76%) | 491 (8.68%) | 90 (19.07%) |

*Note.* NHATS = National Health and Aging Trends Study; SRHD = Self-Reported Hearing Disability; SRVD = Self-Reported Vision Disability. Values for the Cudjoe Social Isolation Measure and the Adjusted Social Isolation Measure ranged from 0 to 5.
<sup>a</sup>Cudjoe et al., 2020.

### Measures

#### Self-Reported Vision Disability

SRVD was measured using three items. Participants were considered to have a vision disability if they reported being blind or if they responded “No” to questions asking if they could see well enough to recognize someone across the street and to read newspaper print while wearing glasses or contact lenses, if applicable. This method has been used in previous studies with NHATS data (Ehrlich et al., 2019; Frank et al., 2019; Xiang et al., 2020).

#### Self-Reported Hearing Disability

SRHD was measured using four items. Participants were coded as having a hearing disability if they reported being deaf or if they responded “No” to questions asking if they could hear well enough to use the telephone, carry on a conversation in a room with the TV or radio playing, and carry on a conversation in a quiet room. This method has been used in previous studies with NHATS data (Kuo et al., 2021). Individuals who self-reported vision disability or hearing disability at baseline were followed longitudinally to better understand trajectories of social isolation.

#### Social Isolation

Preliminary measurement of social isolation was conducted using a five-item composite variable (Cudjoe et al., 2020). Participants received one point for each of the following, for a maximum of five points: if they (a) lived alone, (b) talked to one person or fewer about “important matters” in the past year, (c) did not attend religious services in the past month, (d) did not attend clubs/classes/organized activities in the past month, and (e) did not participate in volunteer work in the past month. Points were summed, with higher scores indicating higher levels of social isolation.

An adjusted measure of social isolation was also constructed, with the goal of identifying elements of familial support which might be more relevant to Hispanic individuals. This adjusted measure did not include the items about participating in volunteer work or attending clubs, etc., but did include items about (a) whether the participant visited with friends/family in the participant’s home or in the home of the friend/family member, and (b) whether they lived in an intergenerational household. Each of these items was included based on familistic values, and because they were theoretically relevant to social connections among Hispanic older adults. The resulting adjusted measure had a maximum of six points, with more points indicating greater social isolation. The original and adjusted measures of social isolation did have considerable overlap, yet also some uniqueness (*r* = .76).

#### Covariates

Covariates included age, gender, and education. Age and education categories were endorsed by participants, and were included in analyses as ordered categorical variables (see Table 1). Participants also reported their gender (0 = male; 1 = female).

### Analytic Approach

We first conducted descriptive analyses of the baseline study sample characteristics (see Table 1). To explore our first research question, unconditional latent growth models were estimated in *Mplus* (Muthén & Muthén, 1998-2017) across two groups (Hispanics and non-Hispanic Whites) using the original measure of social isolation that Cudjoe et al. (2020) described. Group differences in intercept and slope growth curve parameters were tested using the Model Test command in *Mplus*. To address our second research question, steps were taken to explore a more culturally appropriate measurement of social isolation among Hispanic participants. The proportion of each response to binary social isolation items from the original measure described by Cudjoe et al. (2020), as well as some new proposed items, were compared across groups using *z*-tests of proportion (see Table 2). Using the adjusted, more culturally sensitive measure of social isolation that we hypothesized would be more relevant to Hispanic respondents, we estimated multigroup latent growth models of social isolation across time for Hispanic and non-Hispanic White respondents. Group differences in growth curve parameters (i.e., intercepts and slopes) were tested using the Model Test command in *Mplus*. To address the third research question, an additional model included SRVD and SRHD as predictors, along with covariates. Both SRVD and SRHD were examined in relation to the initial levels and slopes of social isolation across time. Education, age, and gender were included as covariates. Finally, sensitivity analyses were conducted to identify possible differences between Hispanic subgroups according to country of origin. Some participants (6% of non-Hispanic White; 8.7% Hispanic) were missing on the social isolation measure at Round 1. The full information maximum likelihood approach was used in estimated models so that all data present from the full sample (N=5,306) could inform estimates.

**Table 2.** Comparison of Frequencies, Percentages, and Z-Tests of Proportions of Social Isolation Responses Between Non-Hispanic White and Hispanic Respondents

| Item | Non-Hispanic<br>Whites | Hispanics | Z-score |
| --- | --- | --- | --- |
|  | Frequency (%) | Frequency (%) |  |
| 1. Participant lives alone. | 1469<br>(30.34%) | 96<br>(21.67%) | 3.83*** |
| 2. Participant has one or fewer people who he/she talked to in the last year about important things. | 2160<br>(46.89%) | 179<br>(45.78%) | .42 |
| 3. In the last month, participant never attended religious services. | 2186<br>(44.98%) | 192<br>(43.15%) | .74 |
| 4. In the last month, {besides religious services,} participant never participated in clubs, classes, or other organized activities.<sup>a</sup> | 3021<br>(62.15%) | 361<br>(81.12%) | -7.97*** |
| 5. In the last month, participant never did volunteer work.<sup>a</sup> | 3609<br>(74.26%) | 408<br>(91.69%) | -8.21*** |
| 6. In the last month, participant did not visit in person with friends or family not living with him/her, either at his/her home or theirs.<sup>b</sup> | 569<br>(11.71%) | 107<br>(24.04%) | -7.47*** |
| 7. Participant does not live in an intergenerational household.<sup>b</sup> | 913<br>(81.21%) | 192<br>(56.85%) | 12.11*** |

<sup>a</sup>Included in the Cudjoe et al. (2020) measure of social isolation; not included in the adjusted Cudjoe et al. (2020) measure.
<sup>b</sup>Included in the adjusted Cudjoe et al. (2020) measure of social isolation; not included in the Cudjoe et al. (2020) measure.
\* $p < .05$ , \*\* $p < .01$ , \*\*\* $p < .001$ .

**Table 3.**
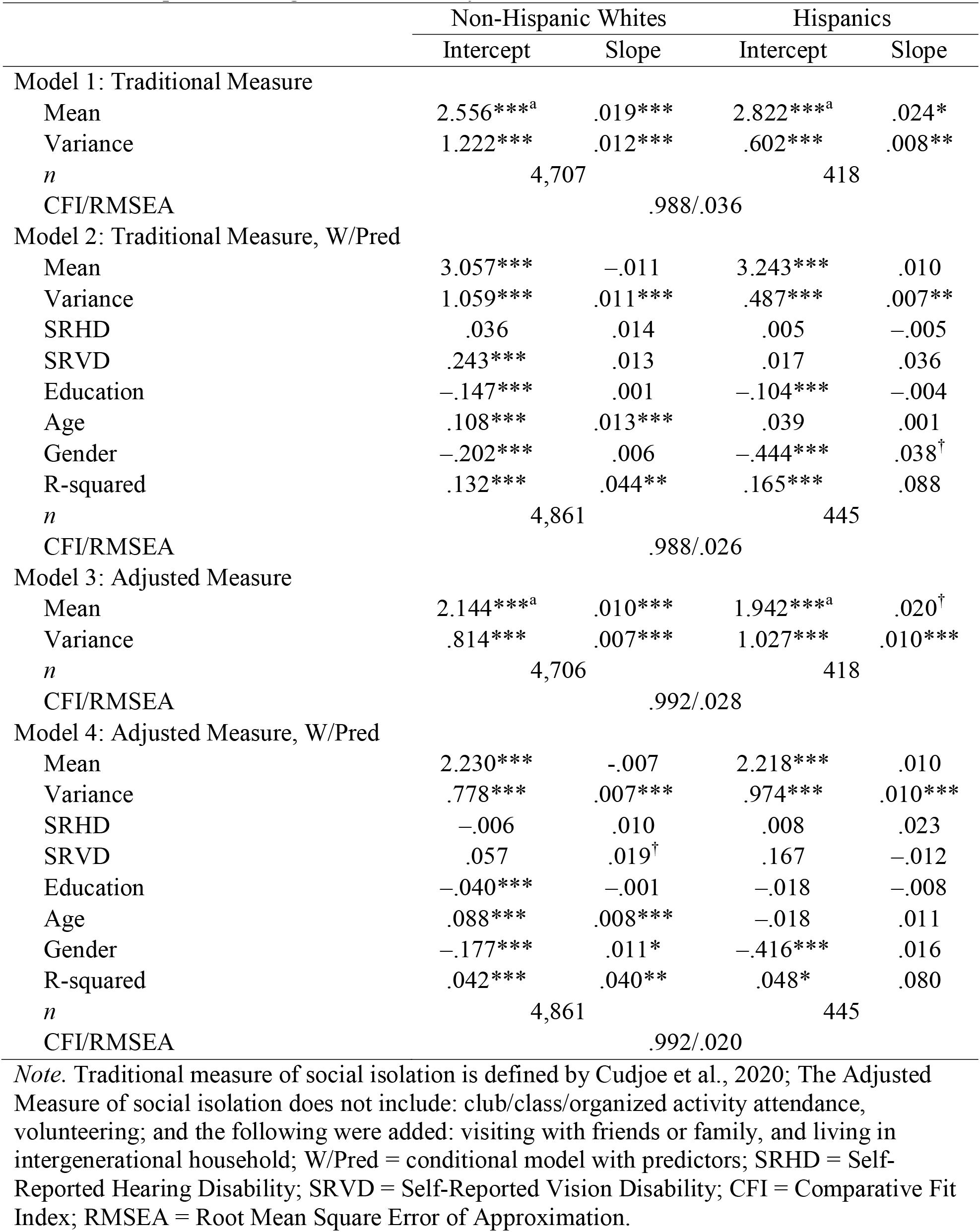
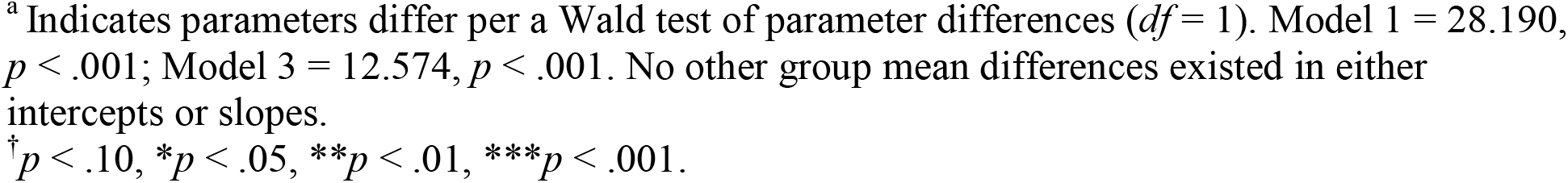
Social Isolation Growth Parameters and Predictors of Those Parameters for Non-Hispanic Whites and Hispanics, Using Two Measures of Social Isolation

|  | Non-Hispanic Whites |  | Hispanics |  |
| --- | --- | --- | --- | --- |
|  | Intercept | Slope | Intercept | Slope |
| Model 1: Traditional Measure |  |  |  |  |
| Mean | 2.556***<sup>a</sup> | .019*** | 2.822***<sup>a</sup> | .024* |
| Variance | 1.222*** | .012*** | .602*** | .008** |
| *n* | 4,707 |  | 418 |  |
| CFI/RMSEA | .988/.036 |  |  |  |
| Model 2: Traditional Measure, W/Pred |  |  |  |  |
| Mean | 3.057*** | −.011 | 3.243*** | .010 |
| Variance | 1.059*** | .011*** | .487*** | .007** |
| SRHD | .036 | .014 | .005 | −.005 |
| SRVD | .243*** | .013 | .017 | .036 |
| Education | −.147*** | .001 | −.104*** | −.004 |
| Age | .108*** | .013*** | .039 | .001 |
| Gender | −.202*** | .006 | −.444*** | .038<sup>†</sup> |
| R-squared | .132*** | .044** | .165*** | .088 |
| *n* | 4,861 |  | 445 |  |
| CFI/RMSEA | .988/.026 |  |  |  |
| Model 3: Adjusted Measure |  |  |  |  |
| Mean | 2.144***<sup>a</sup> | .010*** | 1.942***<sup>a</sup> | .020<sup>†</sup> |
| Variance | .814*** | .007*** | 1.027*** | .010*** |
| *n* | 4,706 |  | 418 |  |
| CFI/RMSEA | .992/.028 |  |  |  |
| Model 4: Adjusted Measure, W/Pred |  |  |  |  |
| Mean | 2.230*** | −.007 | 2.218*** | .010 |
| Variance | .778*** | .007*** | .974*** | .010*** |
| SRHD | −.006 | .010 | .008 | .023 |
| SRVD | .057 | .019<sup>†</sup> | .167 | −.012 |
| Education | −.040*** | −.001 | −.018 | −.008 |
| Age | .088*** | .008*** | −.018 | .011 |
| Gender | −.177*** | .011* | −.416*** | .016 |
| R-squared | .042*** | .040** | .048* | .080 |
| *n* | 4,861 |  | 445 |  |
| CFI/RMSEA | .992/.020 |  |  |  |

*Note.* Traditional measure of social isolation is defined by Cudjoe et al., 2020; The Adjusted Measure of social isolation does not include: club/class/organized activity attendance, volunteering; and the following were added: visiting with friends or family, and living in intergenerational household; W/Pred = conditional model with predictors; SRHD = Self-Reported Hearing Disability; SRVD = Self-Reported Vision Disability; CFI = Comparative Fit Index; RMSEA = Root Mean Square Error of Approximation.
<sup>a</sup> Indicates parameters differ per a Wald test of parameter differences ( $df = 1$ ). Model 1 = 28.190, $p < .001$ ; Model 3 = 12.574, $p < .001$ . No other group mean differences existed in either intercepts or slopes.
<sup>†</sup> $p < .10$ , $*p < .05$ , $**p < .01$ , $***p < .001$ .

## Results

### Preliminary Analyses

Responses to individual items in the social isolation measure were analyzed using *z*-tests of proportion to compare differences between Hispanics and non-Hispanic Whites (see Table 2). Results suggested that Hispanic participants were more likely to live with others and were less likely to visit clubs and participate in formal volunteer opportunities. No group differences were found in talking with others about important things or participating in religious services.

A comparison of responses to the new, culturally adjusted survey questions were also made between Hispanic and non-Hispanic White respondents using z-tests of proportion (see Table 2). Results suggested that Hispanic participants were less likely than non-Hispanic Whites to visit with friends or family in their home or the homes of others, yet were more likely to live in an intergenerational household.

### Social Isolation Trajectories with Traditional Social Isolation Measure (Model 1)

An unconditional growth curve model was estimated to examine the intercepts and slopes of social isolation for non-Hispanic Whites and Hispanics. The growth curve model provided adequate fit to the data (Comparative Fit Index [CFI] = .988; Root Mean Square Error of Approximation [RMSEA] = .036, 90% confidence interval, or CI = [.032, .041]). Initial levels of social isolation differed significantly across groups (non-Hispanic White *M*_intercept_ = 2.556, *p* < .001; Hispanic *M*_intercept_ = 2.822, *p* < .001; Wald value = 28.190 (df = 1), *p* < .001). Slopes of social isolation increased significantly across time among both groups (non-Hispanic White *M*_slope_ = .019, *p* < .001; Hispanic *M*_slope_ = .024, *p* < .05), but did not significantly differ (Wald value = .206 (df = 1), *p* = .65). Panel A of Figure 1 shows a visual representation of social isolation trajectories with the traditional social isolation measure.

### Sensory Disability and Social Isolation Trajectories (Model 2)

SRVD and SRHD were added as predictors of initial levels and slopes of social isolation trajectories for non-Hispanic White and Hispanic participants using the traditional measure of social isolation (Cudjoe et al., 2020) while controlling for all other covariates. Model 2 adequately fit the data (CFI = .988; RMSEA = .026, 90% CI = [.022, .029]). SRVD was associated with higher initial levels of social isolation for non-Hispanic White participants (β [SE] *=* .24 [.069], *p* = .01). No other sensory disability predictors were associated with initial levels or slopes of social isolation.

### Social Isolation Trajectories with Adjusted Social Isolation Measure (Model 3)

Using the novel measure of social isolation, a growth curve of social isolation was estimated to assess the intercepts and slopes for the Hispanic and non-Hispanic White participants. The model provided adequate fit to the data (CFI = .993; RMSEA = .028, 90% CI = [.023, .032]). Non-Hispanic Whites (*M*_intercept_ = 3.017, *p* < .001) and Hispanics (*M*_intercept_ = 2.852, *p* < .001) significantly differed in their initial levels of social isolation (Wald value = 7.925 (df = 1), *p* < .01). Mean slope values suggested significant increases in social isolation across time for both Hispanic (*M*_slope_ = .025, *p* < .05) and non-Hispanic White (*M*_slope_ = .015, *p* <.001) respondents. Panel B of Figure 1 shows a visual representation of social isolation trajectories with the adjusted social isolation measure.

### Sensory Disability and Social Isolation Trajectories with Adjusted Social Isolation Measure (Model 4)

Using the novel social isolation measure, SRVD and SRHD were used as predictors of intercepts and slopes of social isolation trajectories for non-Hispanic White and Hispanic participants while controlling for all other covariates. Model 4 provided adequate fit (CFI = .994; RMSEA = .018, 90% CI = [.014, .022]). Neither SRVD nor SRHD was significantly related to the intercept or slope of social isolation for Hispanic or non-Hispanic White participants.

### Sensitivity Analyses

Sensitivity analyses were performed to examine country of origin as a possible contextual variable that might shape findings regarding SRVD and SRHD as predictors of intercepts and slopes of social isolation trajectories. Six models were assessed (see Supplemental Table 1), including some with the original social isolation measure and some with the adjusted measure. Results from those analyses suggest some differences between Hispanics of various national origins (Mexican, Puerto Rican, and Other) and non-Hispanic Whites in their social isolation trajectories as well as how SRVD and SRHD relate. Among other findings, SRVD is associated with an increased slope of social isolation across time among the Mexican group when the original measure of social isolation was used, but this association was no longer present when the adjusted measure of social isolation was used. This pattern was not observed in groups of Hispanic participants from other national origins.

## Discussion

Given the association between sensory loss and social isolation, as well as evidence of health disparities related to sensory loss between Hispanic and non-Hispanic White older adults (Arnold et al., 2019; Varma et al., 2004; West & Scott, 2021; Uribe, 2011), the purpose of this study was to explore the impact of self-reported sensory disability on social isolation among those of Hispanic ethnicity. We hypothesized that, despite disparities in access to screening and treatment for sensory loss, Hispanic older adults may be less socially isolated than their non-Hispanic White counterparts, due to the influence of the cultural values that promote a reliance on family ties, or familism.

Initial analyses revealed that levels of social isolation were unexpectedly higher among Hispanic than non-Hispanic White participants. This was unanticipated, as past research has described how the family-centered culture among Hispanic individuals serves as a major buffer against stress, social isolation, and other negative health outcomes (Corona et al., 2017; Markides et al., 2019). Analyzing individual items in the composite measure of social isolation proposed by Cudjoe et al. (2020) revealed that, though Hispanic participants initially appeared more socially isolated, they reported that they were less likely to live alone than their non-Hispanic White counterparts. Further analyses revealed that Hispanic respondents were less likely to report participation in community activities outside of the home, including doing volunteer work or attending clubs or classes. This led us to hypothesize that perhaps the existing composite measure did not include features of social support that are important to Hispanic individuals, including family connections. Thus, we created a new, modified indicator of social isolation that included culturally relevant items for Hispanic older adults which emphasized familial support.

When we applied the original Cudjoe et al. (2020) measure of social isolation, Hispanic older adults with sensory disabilities appeared to be significantly more socially isolated than their non-Hispanic white counterparts. Using the new measure of social isolation, which included measures of family support, it appeared that the Hispanic sample were significantly less socially isolated than they originally appeared. The Hispanic respondents in the study were more likely than non-Hispanic Whites to live in intergenerational households and were less likely to live alone. This trend is reflected in other national data. As of 2016, 20% of Americans live in multi-generational households (up from 17% in 2009) and this trend is increasing. Racial and ethnic minorities are more likely to live in multi-generational households (% in 2016 from % in 2009, respectively): Hispanic 27% from 23%; Asian 29% from 26%; Black 26% from 24%; non-Hispanic Whites 16% from 13% (Cohn & Passel, 2018). In the current study, 42% of Hispanic participants reported living in an intergenerational household, which may be evidence of familism at play, as family members may be providing social connection and other support for older parents and grandparents who, in this sample, are living with sensory disabilities. Despite this, Hispanic respondents reported they were less likely to have visitors or to visit others. This could be evidence of perceived social isolation driven by unfulfilled, culturally driven social expectations due to acculturation of younger generations (Markides et al., 2013).

We explored the impact of self-reported sensory disabilities on social isolation and discovered that, when using the traditional measure (Cudjoe et al., 2020), vision disability (SRVD) was significantly associated with higher levels of social isolation for non-Hispanic Whites, but not for Hispanic participants. This is further evidence that cultural factors may be at play and may buffer the impact of sensory disability, even before the new adjusted measure of social isolation is used. Hearing disability (SRHD) was not associated with initial levels or longitudinal increases in social isolation for either group and there were no differences between groups when using the new adjusted measure of social isolation. This could be because these measures are not capturing the types of social isolation someone with a hearing disability may experience, including subjective feelings of disconnection, even while they are participating in events in the community or with family.

### Strengths and Limitations

Approximately 1.2% of undocumented residents of the U.S. are age 65+ (Pew Research Center’s Hispanic Trends Project, 2020). The NHATS data are derived from Medicare beneficiaries and thus may not be representative of individuals not eligible for Medicare. When designing the new, culturally sensitive measure of social isolation, we were limited by the constraints of the data. We could select only the items that were included in the NHATS survey. There are items or measures not included in NHATS that may have been appropriate to assess social activities that are more common among Hispanic individuals such as “attending a dance” or “picnicking on the weekend.” Still, our study represents an important starting point, utilizing a nationally representative sample to generate early evidence that cultural values may be at play, not only in the way research participants describe their level of social isolation, but also in how families respond to their older family members who are living with disabilities, such as sensory disability. Shedding light on these patterns allows us to better understand how to support older adults and their family units as they navigate life with sensory loss.

### Future Directions

Future research, including mixed-methods work, is needed to create culturally sensitive measures of social isolation and measures of other important culturally mediated health outcomes. Such measures could involve greater nuance in response options, and therefore could be subjected to psychometric testing and development. Additionally, future investigations could explore familial and other social dynamics among older adults of Hispanic origin with sensory disabilities. Hispanics are a heterogeneous group comprised of individuals from various countries, historical and political backgrounds and races. Although familism is a shared value among Hispanics, there are likely differences between subgroups due to their heterogeneity, as our preliminary sensitivity analyses suggest (supplemental Table 1). Attention should be given to understanding differences between Hispanic subgroups (e.g., those born in the U.S., those from various countries, or those dealing with challenges related to immigration status in which family structures may be disrupted). This may be facilitated by upcoming NHATS rounds that will include larger, more diverse samples due to planned oversampling of Hispanic participants. In addition, the current NHATS measures of sensory disability represent subjective assessments. In future rounds of NHATS data, where objective measures of vision and hearing impairment are included, research could explore nuances of how degree of vision or hearing disability relate differently to social isolation.

## Conclusion

This investigation suggests that measures of social isolation that focus on social support outside of the home may not be valid among Hispanic older adults. Culturally sensitive measures of social isolation will be increasingly consequential for future research and health policy to meet the needs of a diverse older population.

## Data Availability

Data, analytic methods and study materials will be made available upon request to the authors.

## Acknowledgments

data, analytic methods and study materials will be made available upon request.

